# Muscle cells of sporadic ALS patients secrete neurotoxic vesicles

**DOI:** 10.1101/2021.03.11.21252078

**Authors:** Laura Le Gall, William J Duddy, Cecile Martinat, Virginie Mariot, Owen Connolly, Vanessa Milla, Ekene Anakor, Zamalou G Ouandaogo, Stephanie Millecamps, Jeanne Lainé, Udaya Geetha Vijayakumar, Susan Knoblach, Cedric Raoul, Olivier Lucas, Jean Philippe Loeffler, Peter Bede, Anthony Behin, Helene Blasco, Gaelle Bruneteau, Maria Del Mar Amador, David Devos, Alexandre Henriques, Adele Hesters, Lucette Lacomblez, Pascal Laforet, Timothee Langlet, Pascal Leblanc, Nadine Le Forestier, Thierry Maisonobe, Vincent Meininger, Laura Robelin, Francois Salachas, Tanya Stojkovic, Giorgia Querin, Julie Dumonceaux, Gillian Butler Browne, Jose-Luis González De Aguilar, Stephanie Duguez, Pierre Francois Pradat

**Affiliations:** Northern Ireland Center for Stratified Medicine, Biomedical Sciences Research Institute, Londonderry, UK; Sorbonne Université, Institut National de la Santé et de la Recherche Médicale, Association Institut de Myologie, Centre de Recherche en Myologie, UMRS974, Paris, France; I-Stem, INSERM/UEVE UMR 861, I-STEM, AFM, Corbeil-Essones, France; NIHR Biomedical Research Centre, University College London, Great Ormond Street Institute of Child Health and Great Ormond Street Hospital NHS Trust, London, UK; Inserm U1127, CNRS UMR7225, Sorbonne Universités, UPMC Univ Paris 6 UMRS1127, France; George Washington University, Genetic Medicine, Children’s National Medical Center, Washington, US; The Neuroscience Institute of Montpellier, Inserm UMR1051, Univ Montpellier, Saint Eloi Hospital, Montpellier, France; Mécanismes Centraux et Périphériques de la Neurodégénérescence, Université de Strasbourg, INSERM UMR_S 1118, Strasbourg, France; Computational Neuroimaging Group, Academic Unit of Neurology, Trinity College Dublin, Ireland; Sorbonne Université, CNRS, INSERM, Laboratoire d’Imagerie Biomédicale, Paris, France; APHP, Département de Neurologie, Hôpital Pitié-Salpêtrière, Centre référent SLA, Paris, France; APHP, Centre de référence des maladies neuromusculaires Nord/Est/Ile de France, Institut de Myologie, Hôpital Pitié-Salpêtrière, Paris, France; Laboratoire de Biochimie et Biologie Moléculaire, Hôpital Bretonneau, CHRU de Tours, Tours, France; INSERM U1171, Pharmacologie Médicale & Neurologie Université, Faculté de Médecine, CHU de Lille, Lille, France; Département de Neurologie, Centre de Référence Maladies Neuromusculaires Paris-Est, Hôpital Raymond- Poincaré, Garches, France; Laboratory of Molecular Biology of the Cell, Ecole Normale Supérieure de Lyon, Lyon, France; Hôpital des Peupliers, Ramsay Générale de Santé, F-75013 Paris, France

**Keywords:** Secreted vesicles, Cell-cell communication, MND, sporadic ALS

## Abstract

**Background:** The cause of the motor neuron (MN) death that drives terminal pathology in Amyotrophic Lateral Sclerosis (ALS) remains unknown, and it is thought that the cellular environment of the MN may play a key role in MN survival. Several lines of evidence implicate vesicles in ALS, including that extracellular vesicles may carry toxic elements from astrocytes towards motor neurons, and that pathological proteins have been identified in circulating extracellular vesicles of sporadic ALS patients. Since MN degeneration at the neuromuscular junction is a feature of ALS, and muscle is a vesicle-secretory tissue, we hypothesized that muscle vesicles may be involved in ALS pathology.

**Methods:** Sporadic ALS patients were confirmed to be ALS according to El Escorial criteria, were genotyped to test for classic gene mutations associated with ALS, and physical function was assessed using the ALSFRS-R score. Muscle biopsies of either mildly affected deltoids of ALS patients (n=27) or deltoids of aged-matched healthy subjects (n=30) were used for extraction of muscle stem cells, to perform immunohistology, or for electron microscopy. Muscle stem cells were characterized by immunostaining, RTqPCR and transcriptomic analysis. Secreted muscle vesicles were characterized by proteomic analysis, Western blot, NanoSight, and electron microscopy. The effects of muscle vesicles isolated from the culture medium of ALS and healthy myotubes were tested on healthy human-derived iPSC motor neurons and on healthy human myotubes, with untreated cells used as controls.

**Results:** An accumulation of multivesicular bodies was observed in muscle biopsies of sporadic ALS patients by immunostaining and electron microscopy. Study of muscle biopsies and biopsy-derived denervation-naïve differentiated muscle stem cells (myotubes) revealed a consistent disease signature in ALS myotubes, including intracellular accumulation of exosome-like vesicles and disruption of RNA-processing. Compared to vesicles from healthy control myotubes, when administered to healthy motor neurons the vesicles of ALS myotubes induced shortened, less branched neurites, cell death, and disrupted localization of RNA and RNA-processing proteins. The RNA-processing protein FUS and a majority of its binding partners were present in ALS muscle vesicles, and toxicity was dependent on the expression level of FUS in recipient cells. Toxicity to recipient motor neurons was abolished by anti-CD63 immuno-blocking of vesicle uptake.

**Conclusion:** ALS muscle vesicles are shown to be toxic to motor neurons, which establishes the skeletal muscle as a potential source of vesicle-mediated toxicity in ALS.

**One Sentence Summary:** Muscle cells of ALS patients secrete vesicles that are toxic to motor neurons

## Introduction

Amyotrophic lateral sclerosis (ALS) is a fatal adult-onset motor neuron disorder affecting 3-5/100,000 individuals per year ^1^. The cause of pathology is likely complex with onset resulting from some combination of genetic mutations, DNA damage, environmental risk factors, viral infections, or other factors, leading to diverse cellular dysfunction such as glutamate-mediated excitotoxicity, abnormal protein aggregation, and mitochondrial disorganization and dysfunction contributing to oxidative stress (see ^2,3^ for review).

Not only motor neurons (MN) are affected in ALS, but also glial cells, muscle fibers ^4^ and immune cells ^5^, each of which may participate actively in ALS onset and progression. In a FUS murine model of motor neurone disease, where the FUS mutation is expressed in all tissues except the MN, motor deficits still appear at a late stage of the disease ^6^. In addition, when the human mutated SOD1(hSOD1) is selectively knocked-down in MN and astrocytes of newborn transgenic hSOD1 mice, their life expectancy is prolonged by only 65-70 days, and there is still significant astrogliosis and microglial activation ^7^. These studies support the potential role of “non-cell autonomous” MN death in ALS, which could involve multiple cell types ^8^.

Numerous cell and tissue types, including skeletal muscle, can secrete exosomes and other types of vesicle ^9,10^. Such extracellular vesicles may be involved in cell-cell communication in the central nervous system ^11,12^, where they can carry out intercellular transport of functional proteins, mRNA, miRNA, and lipids, and may have key roles in spreading of proteinopathies ^11,12^ or neurotoxic elements ^13^. For instance, astrocytes extracted from the SOD1 murine model of ALS secrete exosomes that contain hSOD1, and propagate this toxic protein to neighbouring MNs ^14^. The present study sought to determine whether vesicle secretion is affected in muscle cells of ALS subjects, and whether ALS muscle vesicles (MuV) could be toxic when taken up by recipient motor neurons.

## Material and Methods

### Participants and ethical approvals

An open biopsy was performed on deltoid muscles of 27 ALS patients with probable or definite ALS according to the revised El Escorial criteria ^15^, (who attended the Motor Neuron Diseases Center (Pitié Salpétrière, Paris). Genetic analyses were carried out on DNA extracted from blood samples for all ALS patients to screen several ALS related genes: the *C9orf72* hexanucleotide repeat expansion (using gene scan and repeat primed PCR procedures described in^16^), *ATXN2* repeat length ^17^ and the coding regions of *SOD1, TARDBP, FUS, UBQLN2* and *TBK1* (sequences of the used primers are available upon request). Thirty deltoid muscle biopsies from healthy age and gender-matched subjects were obtained from the BTR (Bank of Tissues for Research, a partner in the EU network EuroBioBank) in accordance with European recommendations and French legislation. The main demographic, clinical, and genetic characteristics of the subjects are indicated in Table S1.

The protocols (NCT01984957) and (NCT02360891) were approved by the local Ethical Committee and all subjects signed an informed consent in accordance with institutional guidelines.

### Muscle stem cell extraction and culture

Briefly, muscles biopsies were dissociated mechanically as previously described in ^18^, and plated in proliferation medium [1 volume of M199, 4 volumes of Dulbecco’s modified Eagle’s medium (DMEM), 20% foetal bovine serum (v:v), 25 ug.ml^-1^ Fetuin, 0.5 ng.ml^-1^ bFGF, 5 ng.ml^-1^ EGF, 5 ug.ml-1 Insulin]. The myogenic cell population was enriched using CD56 magnetic beads, and for their myogenicity using anti-desmin antibodies as described before ^18^. A minimum of 80% of the cell population were positive for desmin. After rinsing 3 times the proliferative myoblasts with PBS, and 3 times with DMEM to remove any FBS residual, the human muscle stem cells were differentiated into myotubes by culturing them in DMEM for 3 days. All cell cultures were regularly checked every 3 weeks for mycoplasma test. All experiments were conducted in cells at less than 21 divisions to avoid potential cellular senescence, and thus experimental artefacts.

### Muscle vesicle extraction from culture medium

For each replicate, 7.5.10^6^ primary myoblasts with less than 21 divisions were plated in 225 cm^2^ Flask. After 24h, the cells were rinsed 6 times in DMEM, then differentiated into myotubes for 3 days in DMEM. All cell cultures were checked and negative for mycoplasma. Muscle vesicles were extracted from conditioned media as previously described for large-scale isolation protocol ^19,20^. Briefly the conditioned media were centrifuged at 260 g for 10 min at room temperature, then at 4000 g for 20 min at 4°C to remove any dead cells and cell debris, and finally at 20,000g for 1hr at 4°C to remove microparticles. The subsequent supernatant was then filtered through 0.22µm filter to remove any microparticles leftover. The filtered medium was then mixed with total exosome isolation reagent (Life technologies™; 2:1, v:v), incubated overnight at 4°C, and then centrifuged at 10,000 g at 4°C for 1h. The supernatant was discarded and the pellet containing the MuVs was resuspended and rinsed three times in PBS using 100K MWCO column. The 100 µl MuVs suspensions were kept at −80°C until needed. MuVs were either used for treating cell cultures (iPSC-derived motor neurons, or myotubes; treatments being always compared with untreated cells) or for protein content characterization. MuVs protein was extracted using 8M Urea or NuPAGE buffer and quantified using BCA kit. See Figure S2A and supplemental data. The vesicles extracted using this protocol floated at similar density than when extracted by classic ultracentrifugation, 1.15-1.19g.ml^-1^, with a better yield was observed ^20^. Vesicles were positive for CD63, CD81, CD82, Flotillin, ALIX, and negative for calnexin (Figure 2 and ^20^).

### NanoSight

MuVs pellets were resuspended in 100µl of filtered PBS. The MuV suspension was then diluted 10x in PBS. Size and distribution of MuV secreted by primary muscle cells were evaluated by a NanoSight LM10 instrument (NanoSight) equipped with NTA analytic software (version 2.3 build 2.3.5.0033.7-Beta7). Samples were assessed three times as previously described ^21,22^ at temperature set to 22.5C. The minimum particle size, track length and blur were set to “automatic”.

### Muscle vesicles labelling

MuVs were labelled using PKH26 kit (Sigma-Aldrich®). Briefly, after adding 100 µl of Diluent C to the MuV suspension, 100 µl of 4 µM PKH26 solution were added to the sample. After 5 min of incubation, 1 ml PBS was added and the MuVs were washed using a 100K concentrators, 15,000 g at 4°C for 10 min. The MuVs were washed 3 times in PBS using the 100K concentrators before being mixed with the cell media for treatment. All cell cultures treated with MuVs were compared and normalized to untreated cell cultures.

### Muscle vesicles added to iPSC motor neurons

hiPSC-derived motor neurons were obtained as previously described ^23^. 3,000 MN progenitors differentiated for 9 days were then plated in poly-l-ornithine (SIGMA) Laminin (LifeTechnologies) coated 384 well plates in differentiation medium N2B27 (DMEM F12, Neurobasal v:v, supplemented with N2, B27, Pen-Strep, βMercaptoethanol 0,1%, Glutamax) supplemented with 100nM Rock Inhibitor (RI), 100nM Retinoic Acid (RA), 500nM SAG, 100nM DAPT, 10ng/mL BDNF and Laminin. The medium was replaced at 11 days of differentiation with N2B27 supplemented with 200nM RA, 1µM SAG, 20ng/mL BDNF and 200nM DAPT and again at 14 days of differentiation with N2B27 supplemented with 200nM RA, 20ng/mL GDNF, 20ng/mL BDNF and 200nM DAPT. 16 days differentiated motor neurons were either treated with ALS MuVs or healthy MuVs resuspended in N2B27 differentiation medium supplemented with 20ng/mL GDNF and 20ng/mL BDNF. Cultures were fixed with 4% formaldehyde at 3 days of treatment. The MN were labelled for Tuj1, Islet ½ and analysed as described in the immuno-labelling section. MN loss was normalized to untreated MN cultures. All cell culture treated with MuVs were compared and normalized to untreated cell culture.

### Muscle vesicles pre-treatment with CD63 antibody

After labelling 0.5µg MuVs with PKH26 as described above, the MuV suspension was incubated for 2 hr at RT with 0.5 µg of CD63 antibody (TS63, LifeTechnologies) and then added to the culture medium of hiPSC-derived motor neurons as described in the paragraph “Muscle vesicles added to iPSC motor neurons”. All cell cultures treated with MuVs were compared and normalized to untreated cell cultures.

### Muscle vesicles added to healthy human muscle cells

Labelled MuVs were added to the differentiation medium of 200,000 control cells cultured in Ibidi 35mm µ-Dishes. MuVs absorption occurred during the first 3 days of differentiation. The myotubes were then rinsed 3 times with PBS, and fresh DMEM was added to the petri-dishes. The cells were fixed with 3,6% formaldehyde for 15min at room temperature at day 3 or day 7 of differentiation, then washed 3 times in PBS and stored at 4C until subsequent analysis:

- *Myonuclear domain* - The myotubes were fixed and stained for DAPI and MF20 as described above. The myonuclear domain was calculated using the following formula:

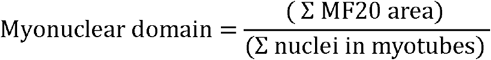
- *Stress blebbing* - Blebs were counted on live images at 4h, 24h, 48h, 72h, 96h and 168h.
- *Cell death inducing an increase in H2Ax expression level* - The myotubes were fixed and stained for DAPI and H2AX as described above. To measure H2AX signal per myonucleus, the total area of H2AX signal was divided by the total number of myonuclei.
- *Cell loss* - The total number of nuclei were counted in each field using ImageJ 1.37v, and summed for each well.
- *Different doses tested* – 4 and 8 µg of MuVs were added to the culture medium of healthy myotubes. H2AX signal per myonucleus, assessed as described above.

All cell cultures treated with MuVs were compared and normalized to untreated cell cultures.

### RNA extraction

Purified muscle stem cells were differentiated for 3 days into myotubes. RNA from muscle cells was extracted as described in ^18^. The quality of RNA samples was assessed with Agilent 2100 Bioanalyzer (Agilent Technologies Inc., Santa Clara, CA).

### Gene expression profiling

- *mRNA gene expression profiling -* Aliquotes of high-quality total RNA from each sample (ALS n=6 and healthy n=6, muscle stem cells) were used for mRNA expression profiling using GeneChip Human Exon 1.0 ST arrays (Affymetrix) as previously described ^24^.
- *Analysis of gene expression data* – see the supplemental data

### Immunolabelling

- *Immunocytochemistry:* 200,000 cells or 100,000 cells were respectively plated on u-dish 35mm high ibidiTreat or 4 wells plate ibidiTreat (ibidi®) in proliferative medium. The following day, muscle stem cells were washed with PBS and myogenic differentiation was induced by cultivating the cells in DMEM only. Muscle cells were fixed at 3 days of differentiation using 4% formaldehyde. The cells were permeabilised, blocked and stained as described ^9^.
- *Immunohistology:* 8 μm muscle transverse sections were cut from human biopsies on a cryostat microtome at −20°C, permeabilised, blocked and stained as previously described ^18^

Primary antibodies used are listed in the Table below and the secondary antibodies used were goat anti-mouse IgG1 or anti-mouse IgG2a, or mouse IgG2b or anti-rabbit tagged with AlexaFluor 355 or AlexaFluor 488 or Alexa Fluor 555 or AlexaFluor 594 or AlexaFluor 647 (1:400, Invitrogen™). The slides were washed, counter-stained with 1µg.ml^-1^ DAPI for 1 min, rinsed 2 times and mounted with ibidi mounting medium (ibidi®).

Ten to 20 non-overlapping pictures were acquired in a line along the diameter of the slide with an Olympus IX70, and an Olympus UPlan FI 10x/0.30 Ph1 and an Olympus BX60 objectives equipped with a Photomatics CoolSNAP™ HQ camera. For the human muscle sections, pictures were taken with an Olympus LCPlan FI 40x/0.60 Ph2 objective. Images were acquired using Zeiss software and analysed using either Fiji or ImageJ 1.37v.

| **Antibody** | **Clone** | **Manufacturer, Catalogue number** | **Species raised, Isotype** | **Dilution** | **Application** |
| --- | --- | --- | --- | --- | --- |
| ALIX | 3A9 | Invitrogen, MA1-83977 | Monoclonal IgG1 | 1:500 | WB |
| Calnexin | AF18 | Life Technologies, MA3-027 | Monoclonal, Mouse IgG1 | 1:100 | WB |
| Caprin (GPIP137) |  | Life Technologies, PA5-29514 | Polyclonal, Rabbit IgG | 1:200 | ICC |
| CD63 |  | BD Pharmingen | Monoclonal,<br>Mouse IgG1 | 1:200 | IHC, ICC |
| CD63 | TS63 | Life Technologies, 10628D | Monoclonal,<br>Mouse IgG1 | 2 µg/ml | WB |
| CD81 | M38 | Invitrogen, 10630D | Monoclonal,<br>Mouse IgG1 | 1:200 | WB |
| CD82 |  | Invitrogen, PA5-27233 | Polyclonal,<br>Rabbit | 1:200 | EM |
| Dystrophin | 3B7 | DSHB, MANDYS1 | Monoclonal,<br>Mouse IgG2a | 1:200 | ICC |
| Flotillin |  | Life Technologies, PA5-18053 | Polyclonal,<br>Goat IgG | 0.3 µg/ml | WB |
| FUS |  | Bethyl, A300-302A | Polyclonal,<br>Rabbit IgG | 1:2000 | ICC |
| FUS |  | Life Technologies, PA5-23696 /PA5-52610 | Polyclonal,<br>Rabbit IgG | 0.4 µg/ml | WB, ICC |
| H2AX |  | Cell Signalling, 763J | Monoclonal,<br>Rabbit IgG | 1:50 | ICC |
| Isl1/2 | 39.4D5 | DSHB | Monoclonal,<br>Mouse IgG2b | 1:100 | ICC |
| Myosin Heavy Chain | MF20 | DSHB | Monoclonal,<br>Mouse IgG2b | 10µg/ml | ICC |
| RPL5 |  | Life Technologies, PA5-26269 / PA5-27539 | Polyclonal,<br>Rabbit IgG | 1:500 | ICC |
| RPL5 |  | Cell Signaling, 14568S | Polyclonal,<br>Rabbit IgG | 1:1000 | WB |
| SOD1 |  | Bethyl, A303-812A | Polyclonal,<br>Rabbit | 0.1 µg/ml | WB |
| TDP43 |  | Invitrogen, PA5-17011 | Polyclonal,<br>Rabbit | 1 :1000 | WB |
| Tsg101 | C-2 | Santa Cruz, sc-7964 | Monoclonal,<br>Mouse IgG2a | 1:200 | ICC |
| Anti-Tubulin Antibody, beta III isoform | TUJ1 | Millipore, MAB-1637 | Monoclonal,<br>Mouse IgG1 | 1:250 | ICC |

### Electron microscopy

- *Electron Microscopy for extracted MuVs -* Purified vesicles were fixed in 2% paraformaldehyde and were counterstained with uranyl and lead citrate and analysed as previously described ^10^.
- *Electron microscopy for human myotubes –* Human myotubes were plated on plastic (Thermanox, Nalge Nunc, Rochester, NY, USA) coverslips and fixed in 2.5% glutaraldehyde in 0.1 M phosphate buffer (v/v), pH 7.4 and further post-fixed in 2% OsO4 (w/v). They were gradually dehydrated in acetone including a 1% uranyl en-bloc staining step in 70% acetone (w/v), and embedded in Epon resin (EMS, Fort Washington, PA, USA). Ultrathin sections were counterstained with uranyl and lead citrate. Observations were made using a CM120 transmission electron microscope (Philips, Eindhoven, The Netherlands) at 80 kV and images recorded with a Morada digital camera (Olympus Soft Imaging Solutions GmbH, Münster, Germany).
- *Electron microscopy for human muscle biopsies –* Human muscles biopsies were fixed in 2% glutaraldehyde, 2% paraformaldehyde, 0.1 M phosphate buffer. After abundant washes and 2% OsO4 post-fixation samples were dehydrated at 4 °C in graded acetone including a 1% uranyl acetate in 70° acetone step and were finally embedded in Epon resin. Thin (70 nm) sections were stained with uranyl acetate and lead citrate, observed using a Philips CM120 electron microscope (Philips Electronics NV) and photographed with a digital SIS Morada camera.

### Analysis of gene expression data

Expression data were uploaded to the GEO repository at accession number GSE122261. Raw data (.CEL Intensity files) were processed using R/Bioconductor packages. Briefly, sample quality was verified by assessment of MA plots, normalized unscaled standard error (NUSE; all samples had median <1.1), and relative log expression (RLE; all sample had divergence <0.2), using the oligo package. Background-corrected normalized log2-transformed probe set signal intensities were obtained by Robust Multi-array Averaging (RMA) using default settings. Affycoretools and huex10stprobeset.db were used to annotate probeset IDs. For gene-level analysis, probesets were retained if they had expression level >log_2_50 in at least 15% of samples. A design matrix (∼0 + condition) was created, to which Limma was used to fit a linear model to the normalized expression values, and differentially expressed genes were identified using Limma’s empirical Bayes method.

For enrichment mapping, the GSEA tool (http://software.broadinstitute.org/gsea/index.jsp) was used to assess the distribution of gene sets across the differential expression profile of ALS compared to Healthy myotubes. 6,349 gene sets were tested, including all of the Gene Ontology Biological Process and Cellular Component collections, and all of the Canonical Pathways from MSigDB. In addition, custom gene sets were created listing genes encoding the known protein binding partners of FUS ^25^ and TDP43 ^26^. Cytoscape v3 and the enrichment map plugin were used to create a graph representing as nodes the gene sets identified by GSEA to be significantly enriched with FDR < 0.05, with edges shown for those pairs of gene sets having overlap coefficient >0.5. Cytoscape’s selection features were used to isolate a sub-graph of the enrichment map showing only the FUS and TDP43 binding gene sets and those gene ontology or canonical pathway gene sets with which they shared genes (overlap coefficient >0.5).

### Quantitative RT-qPCR

- *cDNA synthesis:* RNA from 3 days differentiated muscle cells was extracted as described above. 1 µg of RNA was used to synthesise cDNA using M-MLV Reverse Transcriptase (LifeTechologies™).
- Quantitative PCR was performed on LightCycler® 480 Instrument (Roche) using LightCycler® 480 DNA SYBR Green I Master (Roche).
- *Housekeeping genes*: Beta-2-microglobulin (B2M) showed a constant expression level in all samples and was used to normalize the gene expression levels of extracellular vesicle markers.
- 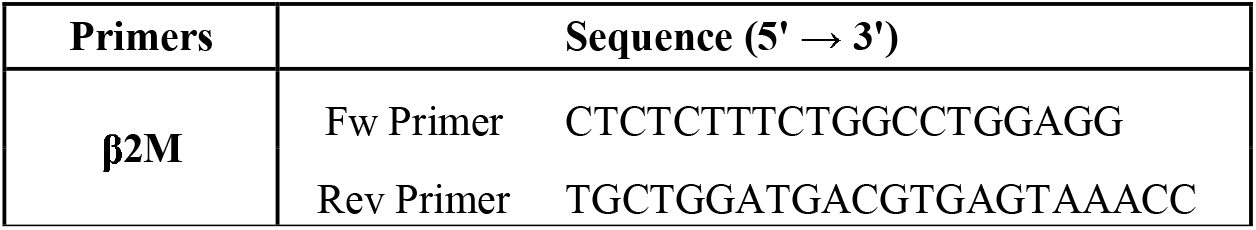
- *Extracellular vesicle markers and FUS expression level:* Quantitative PCR was performed on LightCycler® 480 Instrument (Roche) using LightCycler® 480 DNA SYBR Green I Master (Roche). Primers used are listed in table below. The amplification efficiency of the reaction was calculated using data from a standard curve using RT products from control cells as reference samples (1:20, 1:100, 1:500 and 1:2500 dilutions used). The gene expression level was estimated using the comparative Ct method - Ct representing the cycle at which the fluorescence signal crosses the threshold as previously described ^18^.

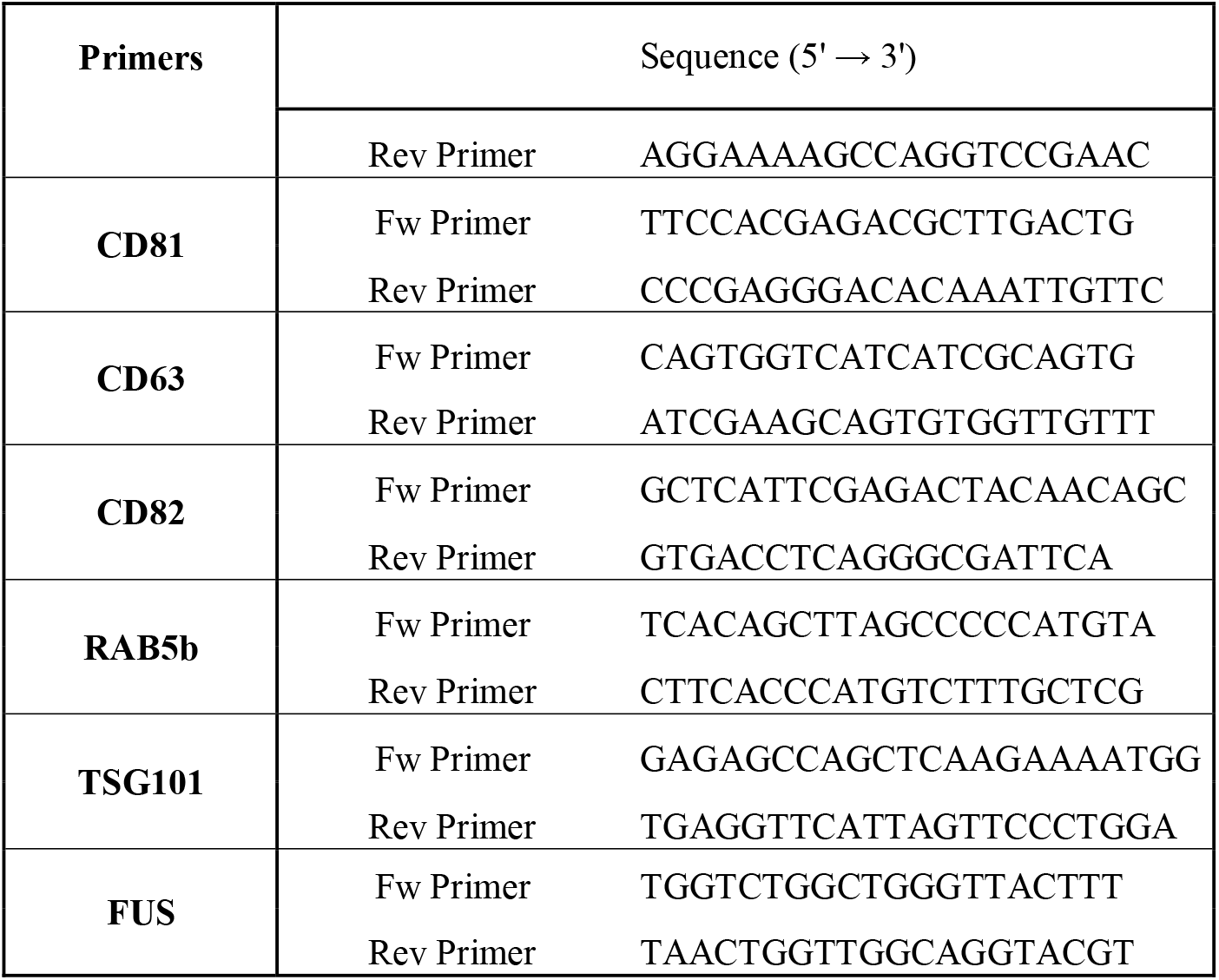

### RNA staining

ALS and healthy muscle cells were differentiated for 3 days before being fixed using 4% paraformaldehyde and permeabilized for 1 hour at RT (5% BSA; 20% FBS; 0,5% Triton X100; 0,5% Tween20). Cells were then stained with a 20 µg/mL acridine orange solution for 10 min at RT.

### Muscle cell lines overexpressing wild type forms of FUS, SOD1 and TDP43

Healthy human muscle cell lines were transduced with previously published plasmid coding either for a tagged form of wild type FUS (FUS-3xFLAG, ^27^), or wild type SOD1 (SOD1-3xFLAG, modified WT-SOD1 plasmid from ^28^) or TDP43 (TDP43-3xFLAG, modified WT-TDP43-YFP plasmid from ^29^) and a selection was done with hygromycin. 50,000 Cont, FUS_FLAG_, TDP43_FLAG_ and SOD1_FLAG_ muscle cells were plated in 8 well Ibidi® plates in proliferative medium. The next day, myogenic differentiation was induced by replacing the medium with DMEM, and cells were treated with 0.5µg of healthy and ALS MuVs. MuVs were applied for 3 days, before fresh DMEM medium was applied to the cultures. At 6 days of differentiation, cells were fixed and analyzed for cell death and cell blebbing as described above. All cell cultures treated with MuV were compared and normalized to untreated cell cultures.

### Proteomic

- The MuV pellets were re-suspended in 25μl 8M Urea, 50 mM ammonium bicarbonate, pH 8.5, and reduced with DTT for 1 h at 4°C. Protein concentrations were then quantified using Pierce BCA Protein Assay kit (ThermoFisher®). MuV proteins were kept at −80°C.
- *Proteome profile determined by Mass spectrometry* - 20 µg of MuV protein were trypsin digested using a SmartDigest column (Thermo) for 2h at 70°C and centrifugated at 1400rpm. Peptides were then fractionated into 8 fractions using a high pH reverse phase spin column (Thermo). Fractioned peptides were vacuum dried, resuspended and analyzed by data-dependent mass spectrometry on a Q Exactive HF (Thermo) with the following parameters: Positive Polarity, m/z 400-2000 MS Resolution 70,000, AGC 3e6, 100ms IT, MS/MS Resolution 17,500, AGC 5e5, 50ms IT, Isolation width 3 m/z, and NCE 30, cycle count 15.
- *Database Search and Quantification* - The MS raw data sets were searched for protein identification for semi tryptic peptides against the Uniprot human database for semi tryptic peptides including common contaminants, using MaxQuant software (version 1.6.2.1) (https://wSww.biochem.mpg.de/5111795/maxquant). We used default parameters for the searches: mass tolerances were set at +/- 20 ppm for first peptide search and +/- 4.5 ppm for main peptide search, maximum two missed cleavage; and the peptide and resulting protein assignments were filtered based on a 1% protein false discovery rate (thus 99% confidence level). 1254 proteins were detected in at least 1 sample. The mass spectrometry proteomics data have been deposited to the ProteomeXchange Consortium via the PRIDE partner repository with the dataset identifier PXD015736.

### Knockdown of FUS with siRNA

100,000 cells were plated in µ-Slide 4 Well ibiTreat (Ibibi®) one day before differentiation was induced. On day 2 of differentiation, cells were transfected using Lipofectamine RNAiMAX Reagent with 200nM s5401 FUS siRNA (LifeTechnologies™) and treated with a low dose of PKH26-labelled ALS MuVs. MuVs were integrated by the cells for 3 days before fresh differentiation medium was added to the 5 days differentiated cells. The cells were harvested at day 8 of differentiation to check for RNA levels by RT-qPCR and to perform cell death analysis, immunostaining for RPL5, and distribution analysis of RNA/DNA using acridine orange staining.

### Western Blotting

Extracted ALS and healthy vesicles were resuspended in lysis buffer (8M urea; 2% SDS; 10 µL/mL protease inhibitor cocktail). Extracted proteins were loaded into NuPage polyacrylamide 4-12% BisTris gels for electrophoresis under reducing conditions (Calnexin, Flotillin, ALIX, FUS, SOD1, TDP43, RPL5) and non-reducing conditions (CD63, CD81). Transfer on polyvinylidene difluoride (PVDF) membrane was performed using the iBlot® Dry Blotting System (Life Technologies™) and upon transfer polyacrylamide gels were stained with Blue Coomassie Gel Code Blue Stain Reagent (LifeTechnologies™) to visualize proteins. Immunoblotting was carried out using the iBind™ Flex Western System and primary antibodies (see Table S3) with respective secondary antibodies (Goat anti Mouse HRP, Donkey anti Goat HRP, Goat anti Rabbit HRP). The signal was detected using the Amersham ECL™ Prime Western blotting Detection Reagent and the UVP ChemiDoc-It^2^ Imager.

### Statistics

All values are presented as means ± SEM. Student’s T-Test was used to compare differences between ALS and control samples for all the protein quantifications, electron microscopy quantifications, MuV integration, immunostaining (FUS, Acridine Orange, RPL5, Caprin1), FUS expression level, MuV-treated MN and myotube death, myotube atrophy. A Kolmogorov-Smirnov test was used to compare the distribution of number of vesicles per MVBs in ALS and healthy myotubes, the distribution of neurites branching in ALS and healthy MuV-treated MN, the distribution of H2AX expression levels in myonuclei treated with different doses of MuVs, and the distribution of myotube nuclear numbers in control myotubes treated with ALS and control MuVs. One-way ANOVA followed by a Tukey’s multiple comparison test was used to evaluate the differences in the in silico secretome, in the immunostaining for extracellular vesicle markers, RT-qPCR, neurites length, and MN treatments. Two-way ANOVA followed by Bonferroni post-hoc test was used to evaluate dose response of MuV on MN survival, changes in bleb counts, and cell death in different cells lines (Cont, FUS_FLAG_, SOD1_FLAG_, TDP43_FLAG_). Enrichment testing p-values were determined by the Fisher’s Exact test (for enrichment testing of custom lists of FUS- and TDP43-binding proteins) or represent a multiple-testing adjusted value based on deviation from expected rank as calculated by the EnrichR tool (for enrichment testing across all GO Molecular Functions). Differences were considered to be statistically different at *P* < 0.05.

## Results

### ALS patient muscle cells accumulate vesicles

To investigate the role of vesicles secreted by ALS muscle cells, myoblasts were extracted from biopsies of confirmed ALS patients at an early stage of pathology (see Table S1 for patient description, and a breakdown of which samples were used in which experiment). Among the 27 sporadic ALS subjects, genetic screening identified only 4 with known ALS-causative mutations (3 carrying the *C9orf72* hexanucleotide repeat expansion, one with aberrant *ATXN2* repeat length). A consistent accumulation of extracellular vesicle markers CD63 (Figure 1A and B) and TSG101 (Figure S1A) in ALS myotubes was observed by immunostaining. Extracellular vesicle markers were higher by RT-qPCR (Figure 1C), and multivesicular bodies filled with exosome-like vesicles were observed by electron microscopy (Figure 1D). *In vivo*, ALS patient muscle biopsies presented an increased frequency of multivesicular bodies (MVBs) in ALS muscles (0.017 MVBs/sarcomere, vs 0.004 MVBs/sarcomere in healthy controls; FigureS1B) and an accumulation of extracellular vesicle markers at the periphery of the myofibers (Figure1E and F).

**Figure 1:**
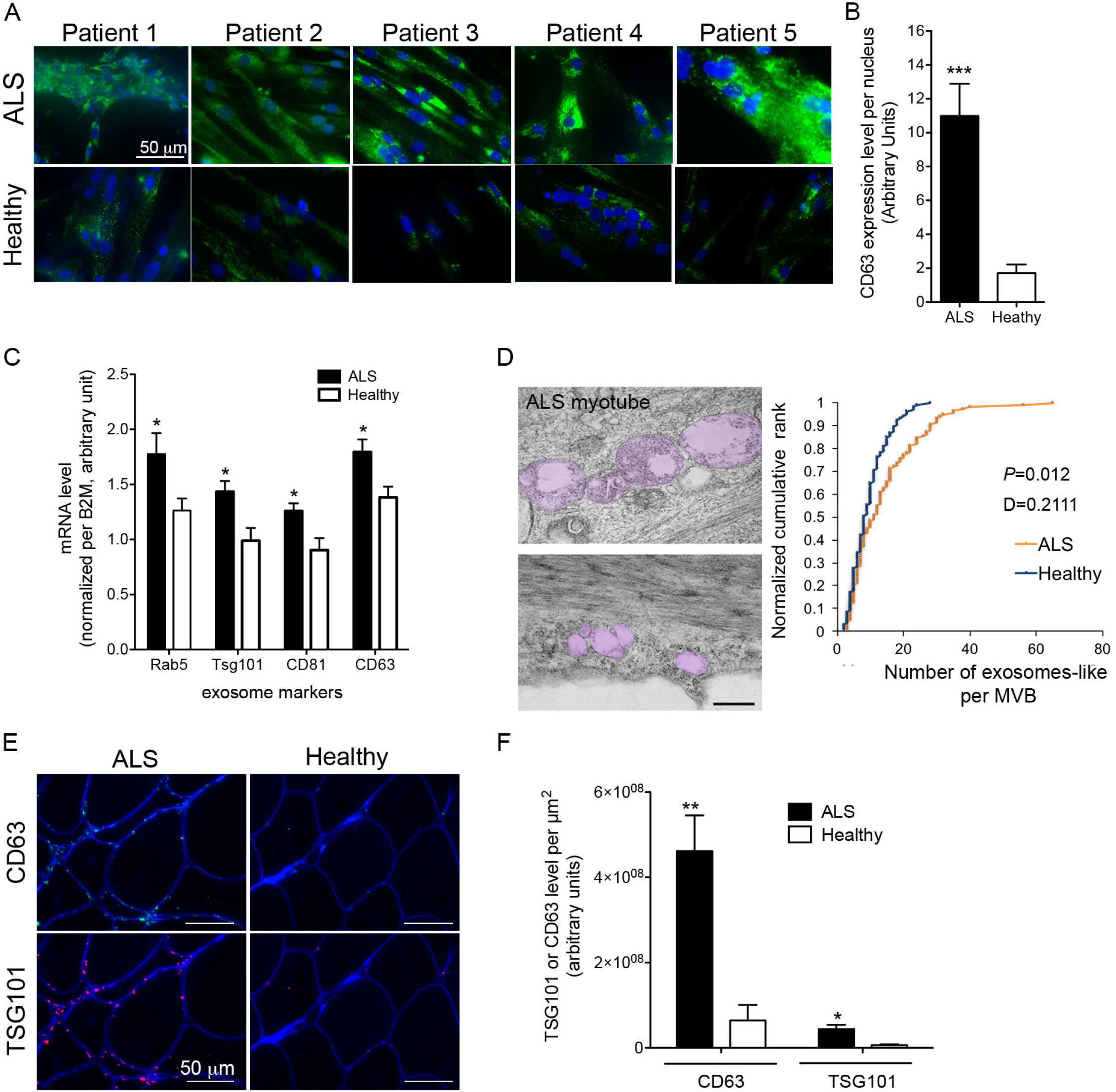
Accumulation of extracellular vesicle markers in myotubes and muscle of ALS patients. (A) Panel showing representative images of CD63 immunostaining performed on cultured myotubes from different patients. (B) Quantification of CD63 fluorescence signal normalized per myonucleus (70-108 myonuclei were analysed per individual, with n=5 ALS and 6 healthy subjects). ***, significantly different from healthy with P<0.001. (C) mRNA encoding for extracellular vesicle proteins normalized per B2M mRNA level are upregulated in ALS myotubes compared to healthy (n=7 ALS, 6 Healthy). * significantly different from Healthy with P<0.05. (D) Multi-vesicular bodies that are present in ALS myotubes contain more exosome-like vesicles than those of healthy controls. Left panel: Representative electron micrographs showing an accumulation of exosome-like vesicles in multi-vesicular bodies (MVBs; highlighted in pink). Scale bar = 500 nm. Right panel: quantification of the number of exosome-like vesicles in the MVBs (100 MVBs per condition were analysed). Two sample Kolmogorov-Smirnov test confirmed the visual impression that the MVBs of ALS muscle contain a greater number of exosome-like vesicles compared to healthy controls (P<0.05). (E) Representative images of extracellular vesicle markers CD63 and TSG101 immunostaining in ALS and healthy muscle. CD63 green, TSG101 red, Dystrophin blue. Scale bar 50 mm. (F) Quantification of extracellular vesicle markers CD63 and TSG101 in muscle biopsies (Pixel per mm^2^, n= 6 ALS, 5 Healthy for CD63, and n=4 per group for TSG101; * and ** significantly different from healthy subjects, *P*<0.05 and *P*<0.01, respectively. Values are means ± SEM. See also Figure S1.

### Secretion of ALS patient muscle cell vesicles

Importantly, since primary muscle stem cells have a limited number of divisions (∼30), all experiments were carried out before the cells reached 21 divisions, thereby avoiding excessive population doublings which could lead to senescence and experimental artefacts ^18,20,30^. In both ALS and healthy vesicle extracts, exosome-like vesicles with a typical cup-shaped morphology (Figure 2A) and sizes ranging from 90-to 200nm (Figure 2A-C) were observed. The muscle vesicles (MuVs) were positive for exosomal markers such as CD63, CD82, CD81, Flotillin, and ALIX, and were negative for calnexin, an endoplasmic reticulum marker normally absent in the exosome fraction (Figure 2C and D). In addition, the vesicle extracts were free of albumin and other contaminants (Figure S2B, and proteomic data GSE122261). The MuV fraction secreted by ALS myotubes contained 1.7-fold more protein than that secreted by an equal number of healthy control myotubes (Figure 2E, Figure S2B).

**Figure 2:**
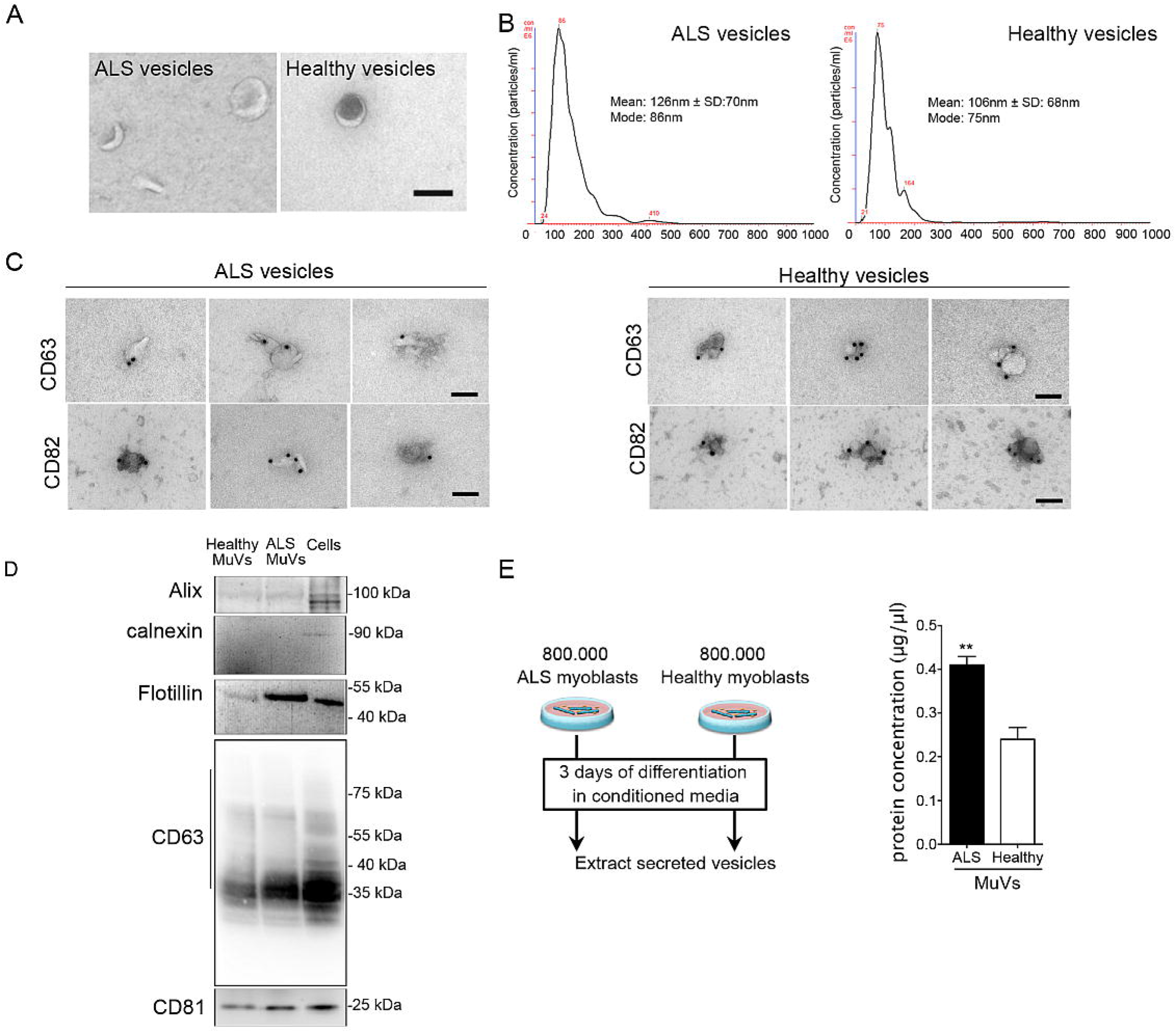
Increased accumulation and secretion of vesicles by ALS myotubes. (A) Representative electron micrographs of vesicles extracted from the culture medium of ALS or healthy myotubes. Scale bar = 100 nm. The extracted vesicles have the typical cup-shape of exosomes. (B) NanoSight analysis showing that ALS and healthy vesicle sizes range between 90-200 nm. (C) Representative electron micrographs of vesicle immunostaining showing that both ALS and healthy muscle vesicles (MuVs) express CD63 and CD82. bar = 100 nm. (D) ALS and healthy muscle vesicles are positive for Alix, flotillin, CD63 and CD81, and negative for calnexin. (E) The MuV fraction secreted by ALS myotubes contained 1.7 more protein than healthy MuV fraction. Left panel: schema summarizing the experimental procedure. Briefly the same number of ALS and healthy myoblasts were differentiated, and after 3 days the culture medium was harvested to extract the MuVs as described in material and methods. Right panel: Protein quantification of MuVs. 800,000 differentiated myoblasts per subject, with n=4 subjects per group. ** significantly different from healthy myotubes (P<0.01). Values are means ± SEM. See also Figure S2.

### Secreted ALS vesicles are neurotoxic

To test neurotoxicity, 0.5 µg of ALS or healthy MuVs were added to the culture medium of healthy human iPSC-derived motor neurons (hiPSC-MN). Since ALS myotubes secrete more MuVs than healthy myotubes, this quantity corresponded to 2 ALS myonuclei or 3.7 healthy myonuclei per MN. Following uptake of MuVs (Figure 3A), only ALS MuVs and not healthy MuVs resulted in shorter neurites (Figure 3B), with less branching (Figure 3C) and a greater cell death (Figure 3D-E) 72h post-treatment. Cell death as a result of ALS MuVs was consistent across multiple patients (Figure 3E), and showed a dose response when different quantities of ALS MuV protein were loaded - ALS MuV toxicity was significant at all concentrations (Figure 3F). Conversely, when MuV uptake was decreased by preincubating MuVs with CD63 antibody (Figure S3A), hiPSC-MN death was dramatically decreased (Figure 3G). Similarly, when added to the culture medium of healthy human myotubes, ALS MuVs induced myotube atrophy (Figure S3B), and cell stress (Figure S3C) leading to cell death (Figure S3D-F). The quantity of cell death was decreased when less MuV protein was loaded, though ALS MuV toxicity remained greater than healthy MuV toxicity at either dose (Figure S3F), suggesting that toxicity is dependent on both the quantity and content of MuVs.

**Figure 3:**
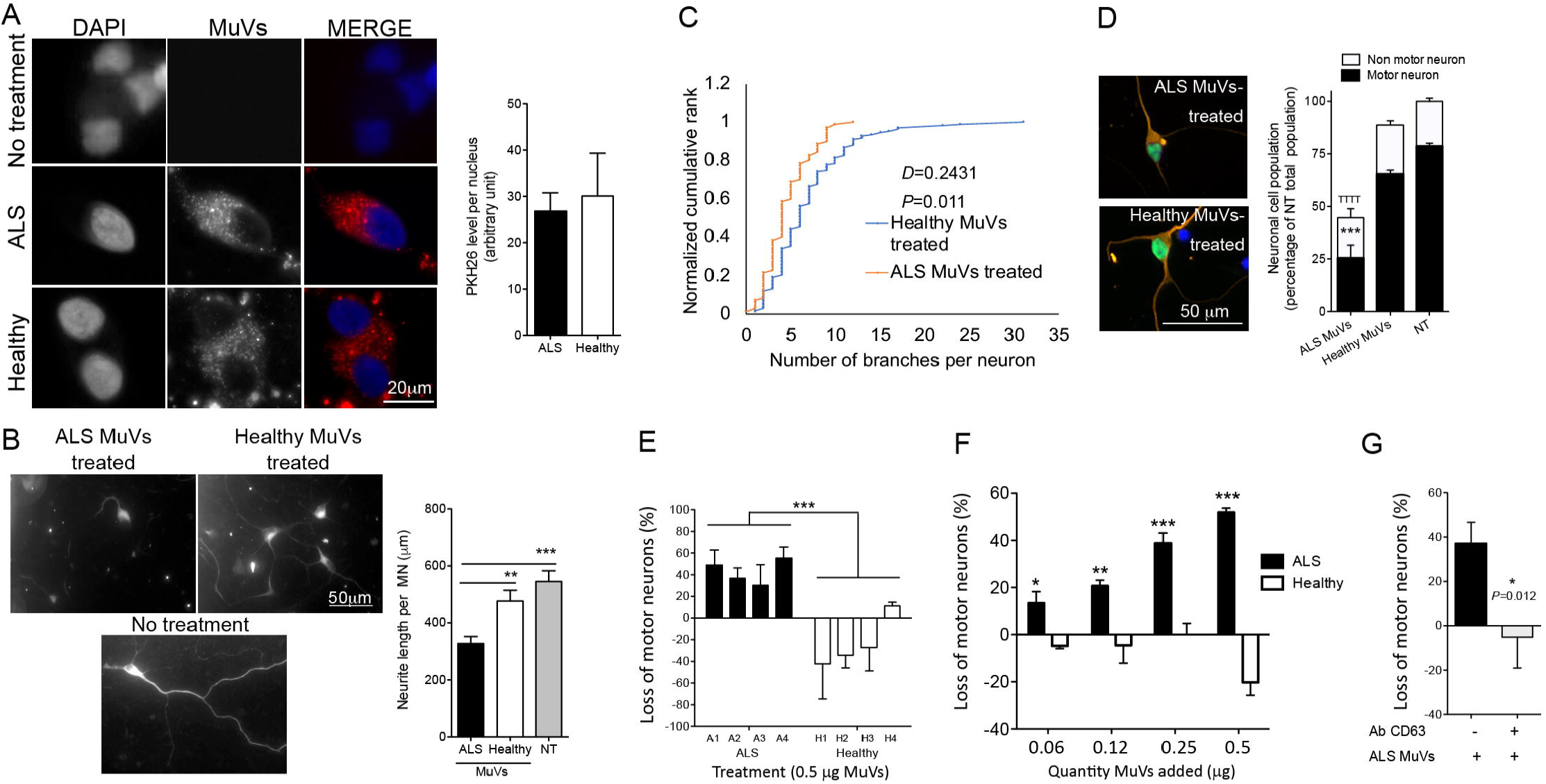
ALS muscle vesicles induce decreased neurite length and branching, and increased death, of human induced pluripotent stem cells derived motor neurons. (A) Equal quantities of ALS and healthy MuVs are taken up by motor neurons. Red: MuVs labelled with PKH26 and added to the culture medium of iPSC-derived motor neurons. (B) Neurite lengths of motor neurons are shortened when treated with ALS MuVs compared to healthy MuVs. Left panel: representative images of MuV-treated motor neurons. Right panel: quantification of neurite length (8 to 15 motor neurons analysed per well, with n=5 wells per treatment). ** and *** significantly different from ALS values *P*<0.01 and *P*<0.001, respectively. (C) Neurites of iPSC-MN cells have fewer neurite branch-points following treatment with ALS MuVs compared to treatment with healthy MuVs; two-sample Kolmogorov-Smirnov test confirmed the visual impression that the number of branches per neurite is decreased (*P*=0.011). (D) Neurotoxicity of ALS MuVs is specific to motor neurons. Right panel: representative images of human iPSC-MN treated with ALS or healthy MuVs. iPSC-MN are positive for motor neuron marker Islet1/2 (green) and neuronal marker Tuj1 (orange). Right panel: quantification of human iPSC neuron and MN cells (10 frames per well, n=3 wells per condition). ***, *P*<0.001 and ^TTT^, *P*<0.001, significantly different from healthy-MuV-treated and non-treated cells, respectively. (E) Quantification of the death of human IPSC-motor neurons treated with equal amounts (by protein content: 0.5 µg) of ALS or healthy MuVs. MuVs were extracted from sporadic ALS patients negative for known mutations (each bar represents a different subject: n=4 subjects per group, each in triplicate). *** P<0.001, significantly different from healthy values. (F) Motor neuron survival in response to increasing concentrations of muscle vesicles from the muscle cells of ALS or healthy subjects. MuVs from the muscle cells of ALS patients (black bars) or healthy subjects (white bars) were added to cultures of iPSC-derived MN at concentrations of 0.06, 0.12, 0.25 or 0.5 µg / 150 µl. The total number of neurons at 72 h after treatment was counted in each condition and expressed as a percentage of the cell death that was observed in cultures of untreated motor neurons at the same time-point. ANOVA 2-factor followed by Bonferroni post-hoc test was performed. * *P*<0.05, ** *P*<0.01, ***, and *P*<0.001, significantly different from healthy-MuV-treated at that concentration. (G) Pre-treatment of the ALS MuVs with CD63 antibody significantly decreased iPSC-MN cell death. * significantly different from ALS values, *P<0*.*05*. Values are means ± SEM. See also Figure S3.

### ALS muscle vesicles are enriched in proteins involved in RNA processing

Proteomic analysis of vesicle content revealed that, of 53 peptides observed at consistently higher levels in the MuVs of ALS subjects compared to Healthy controls, 21 were annotated to the RNA-binding molecular function (Fig.4A; enrichment FDR *P* < 1×10^−7^; Supplementary Table S2). Similarly, of 453 proteins detected only in ALS and not in Healthy controls, 87 were involved in RNA-binding (enrichment FDR *P* < 1×10^−14^; Supplementary Table S3). ALS MuVs contained many known protein binding partners of the RNA-processing proteins FUS ^25^ and TDP43 ^26^ (enrichment *P* < 1×10^−6^; Figure 4B), with 58% (64 of 109) of FUS binding partners being detected, along with FUS itself. FUS and RPL5 – a FUS binding partner – were at a higher level in ALS MuVs by Western blotting (Figure 4C-E). Interestingly, neither SOD1 nor TDP43 were detectable in human MuVs (FigureS4).

**Figure 4:**
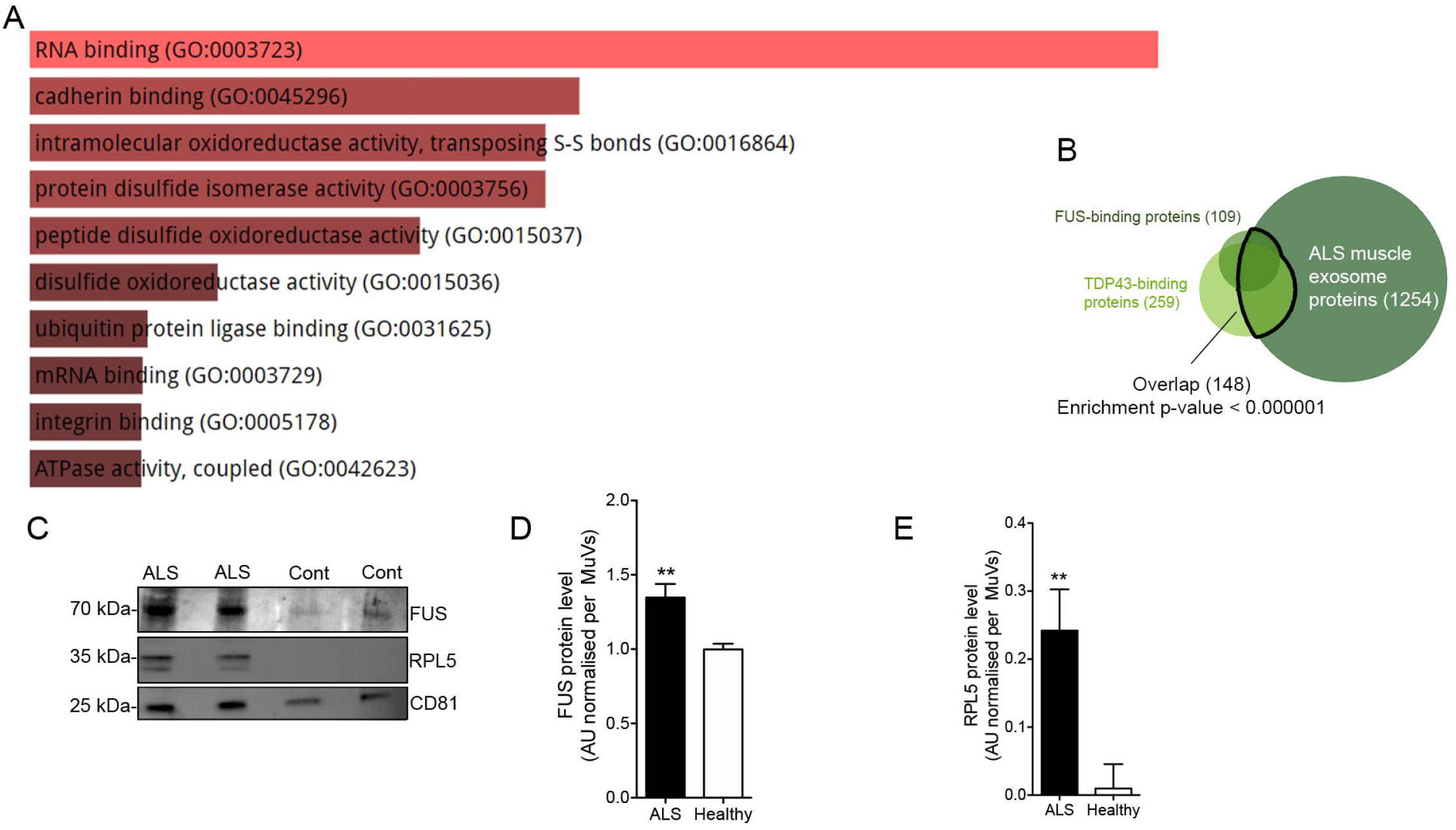
The proteomic content of muscle vesicles is enriched for FUS and TDP43 binding proteins. (A) Output of EnrichR tool showing relative enrichment scores of Gene Ontology Molecular Functions among peptides that were observed at consistently higher levels in the MuVs of ALS subjects compared to Healthy controls (FDR *P*-value for the RNA-binding GO term was < 1×10-7). (B) Whereas only 1.5% of all proteins are known binding partners of FUS and/or TDP43, they represent 13.4% (148 of 1254) of the proteins detected by proteomic analysis of ALS MuVs contents (Fisher’s test P<0.000001). (C) Representative images of Western blot showing the presence of FUS and RPL5 in muscle vesicles. CD81, extracellular vescile markers. (D) FUS protein level in higher in ALS MuVs. Quantification by Western blot of FUS level in MuVs. ** P<0.01, significantly different from healthy values. n=15 ALS and 13 Healthy. (E) RPL5 protein level in higher in ALS MuVs. Quantification by Western blot of RPL5 level in MuVs. * P<0.05, significantly different from healthy values. n=6 subjects per group. Values are means ± SEM. See also Figure S4, Table S2 and S3.

Transcriptomic analysis of cultured myotubes suggested that genes encoding FUS and TDP43 binding proteins were upregulated in the myotubes of ALS patients and that these genes were shared with many RNA-processing pathways that were similarly upregulated (Figure 5A). ALS myotubes presented a greater nuclear accumulation of RNA (Figure 5B), and mis-localization of two FUS protein binding partners, RPL5 and caprin 1, that are involved in RNA processing and stress granule formation (Figure 5C-D) ^25^.

**Figure 5:**
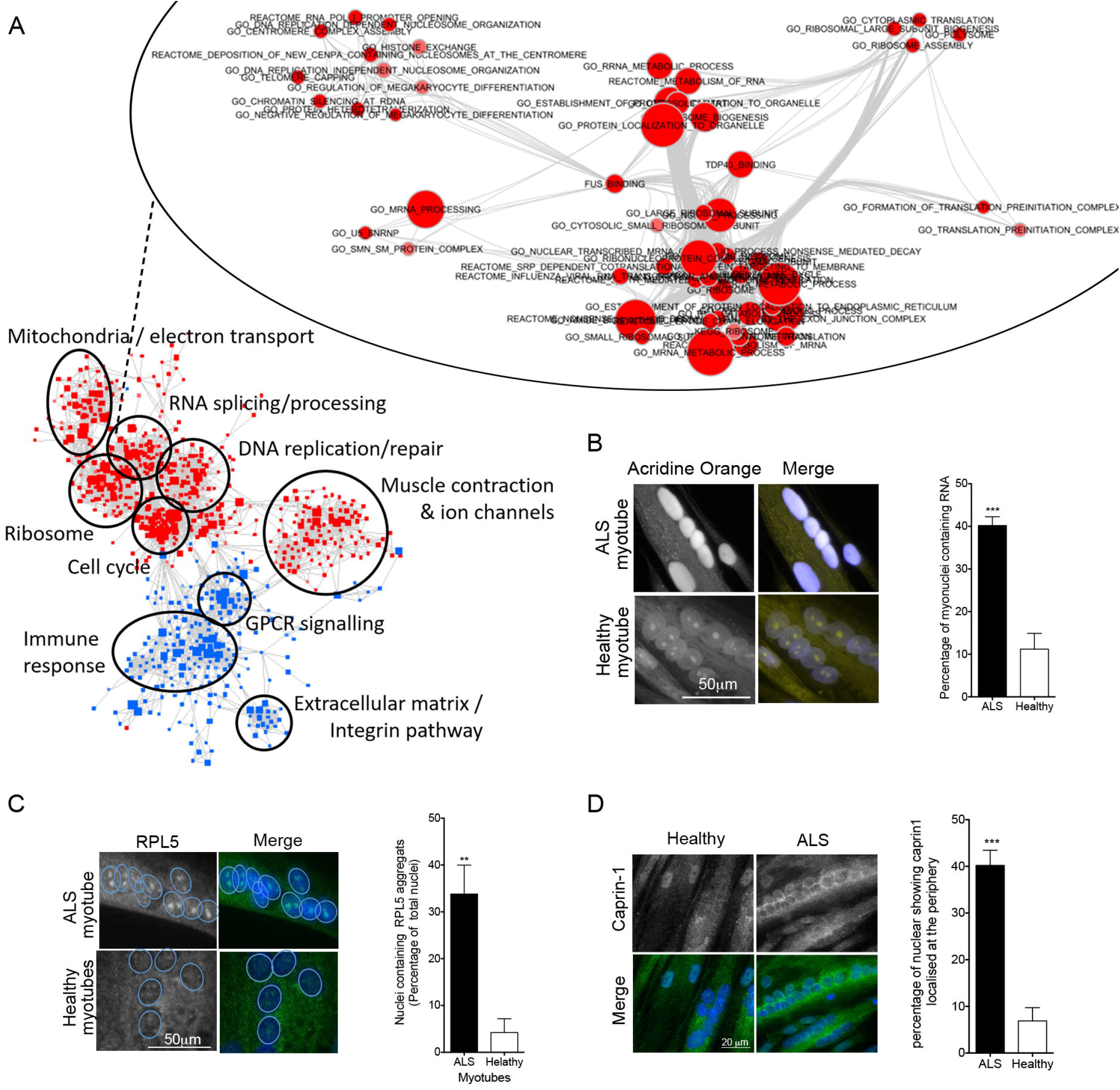
RNA processing is disrupted in ALS myotubes. (A) Enrichment map representing overlap and clustering of the cellular processes and pathways for which gene expression is dysregulated in ALS myotubes compared to healthy controls (red = upregulated; blue = downregulated). Upper inset: Enrichment map showing that genes encoding FUS and TDP43 binding proteins are upregulated in ALS myotubes compared to healthy controls, and that these genes are shared with many RNA-processing pathways that are similarly upregulated (red = upregulated). (B) ALS myotubes present an accumulation of RNA in their nuclei. Left panel: representative images of RNA localisation in ALS and healthy myotubes. Right panel: percentage of myonuclei with high levels of RNA, assayed by acridine orange staining (50 myonuclei analysed per subject, with n=5 subjects per group). ***, *P*<0.001 significantly different from healthy myotubes. (C) RPL5, a protein involved in RNA transport and stress granules, is granulated in ALS myonuclei. Left panel: representative images of RPL5 localisation in ALS and healthy myotubes. Right panel: percentage of myonuclei with RPL5 granules (at least 500 nuclei analysed per subject, with n=4 subjects per group). **, *P*<0.01 significantly different from healthy myotubes. (D) ALS MuVs induce an accumulation of RNA in MN nuclei. Left panel: representative images of RNA localisation in MN nuclei. Right panel: percentage of nuclei with accumulations of RNA (150 to 260 nuclei analysed per well, with n=3 wells per condition). ***, *P*<0.001 significantly different from healthy myotubes. Values are means ± SEM.

### Secreted ALS vesicles affect RNA processing in recipient motor neurons

Since a leading theory is that disruption of RNA metabolism - including RNA translation, transport, storage, and degradation - contributes to ALS physiopathology by affecting neuronal function and viability ^31^, and based on the results described above, we hypothesized an involvement of RNA processing in ALS MuV toxicity. When human iPSC-MNs derived from healthy subjects were treated with ALS MuVs, RNA accumulated in their nuclei (Figure 6A), which has been reported to induce cell death ^32^. Similar results were obtained when ALS MuVs were added to the cultures of human myotubes from healthy subjects (Figure S5A). We hypothesised that ALS MuV toxicity in recipient cells may be mediated through the FUS pathway. To test this, 0.5 µg of ALS or healthy MuVs were added to the culture medium of a human muscle cell line over-expressing a tagged form of wild-type FUS (FUS-_FLAG_; previously published ^27^). This induced an increase in cell death from 8% to 42% (Figure 6B), accompanied with greater cellular stress (Figure S5B) – the same was not observed in cell lines that over-expressed tagged forms of wild-type TDP43 and SOD1 (Figure 6B and S5B). Conversely, when ALS MuVs were added to the culture medium of cells in which FUS was knocked down, lower proportions of nuclei with accumulated RNA (Figure 6C) and RPL5 granules (Figure 6D) were observed, and the quantity of cell death was reduced (Figure 6E). These data implicate the FUS pathway in MuV toxicity and suggest that increased levels of FUS heighten sensitivity to this toxicity, while lower levels reduce it. We note that, relative to muscle cells, high levels of FUS are observed in hiPSC-MN (Figure 6F).

**Figure 6:**
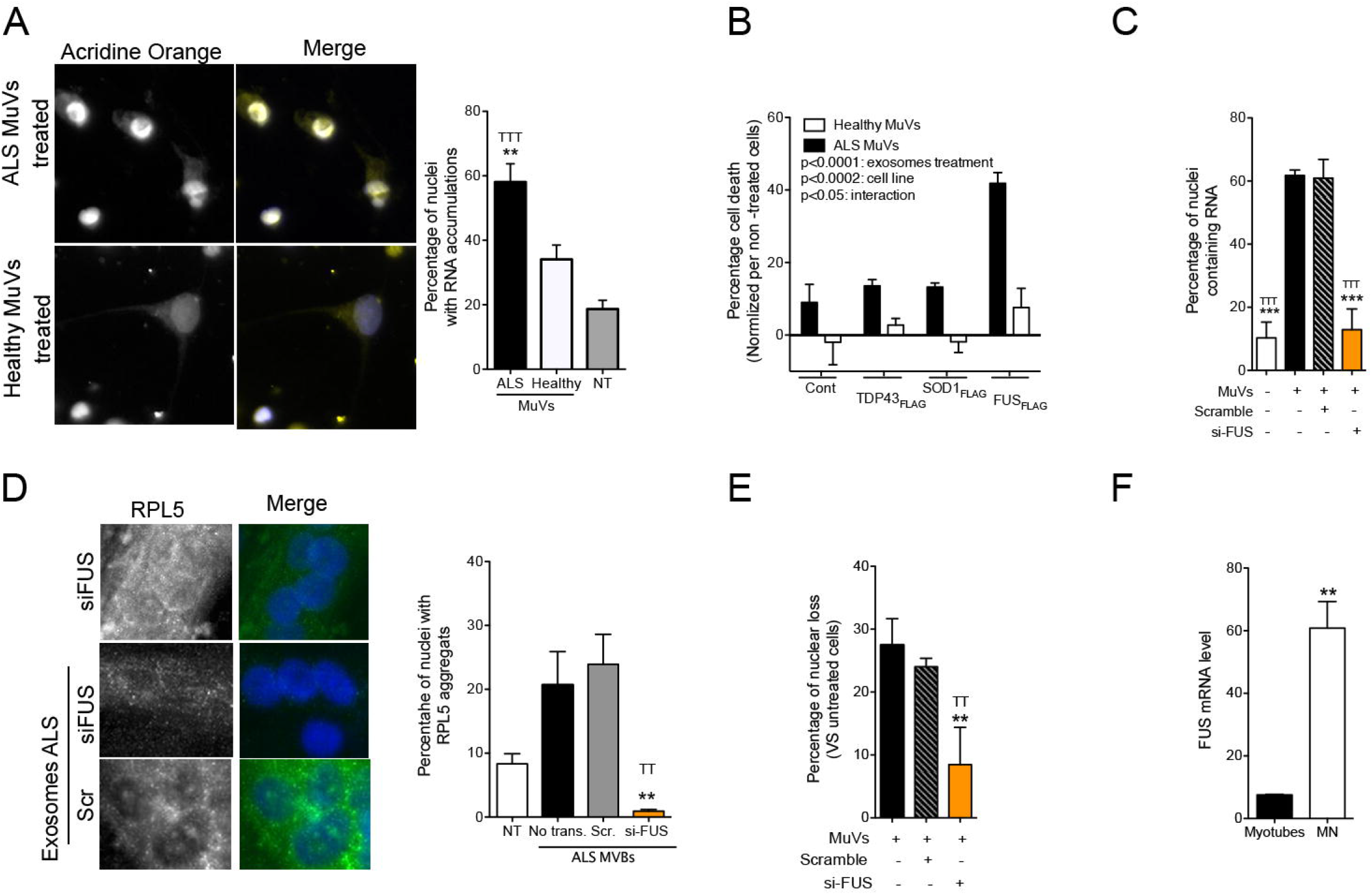
The toxicity of ALS muscle vesicles to healthy motor neurons involves the FUS protein and RNA processing. (A) ALS muscle vesicles induce an accumulation of RNA in MN nuclei. Left panel: representative images of RNA localisation in MN nuclei. Right panel: percentage of nuclei with accumulations of RNA (150 to 260 nuclei analysed per well, with n=3 wells per condition). **, *P*<0.01 and ^TTT^, *P*<0.001 significantly different from MN treated with healthy MuVs and untreated MN, respectively. (B) Cell death induced by 0.5 µg of ALS MuVs is exacerbated in the presence of over-expression of FUS. ANOVA 2 factors was performed, showing the effect of ALS MuVs (P<0.0001), the effect varying due to cell line (*P*<0.0002), and an interaction between the two parameters (P<0.05) (10 frames per well analysed, with n=3 wells per condition). (C) Percentage of nuclei with RNA accumulation are decreased when ALS MuVs are added to the culture medium of cells deficient for FUS (FUS expression level was reduced by 79.5% ± 4.2% with siRNA strategy, 10 frames per well analysed, with n=3 wells per condition). ***, P<0.001 ALS MuV treated cells. ^TTT^, P<0.001 ALS MuV treated scramble-RNA cells respectively. (D) RPL5-aggregates are no longer observed following ALS-MuV-treatment when the recipient cells do not express FUS. Left panel: Representative images of myotubes treated with siFUS or siScrambled control, and with addition or not of ALS MuVs. Myotubes are immunostained for RPL5 (green). Right panel: Percentage of myotube nuclei with RPL5 aggregates in untreated (‘No exo.’), ALS-MuV-treated with no knockdown (‘No trans’), ALS-MuV-treated with siScrambled knockdown (‘Scr.’), or ALS-MuV-treated with siFUS knockdown (‘si-FUS’). Eight hundred to 1,000 nuclei were analysed per well, with n=3 wells per condition. ANOVA 1 factor was performed, showing the effect of ALS MuVs on the two different knock-down conditions, with ** and ^TT^ representing significant difference from ‘No trans.’ and ‘Scr.’, respectively, P<0.01. (E) Percentage of cell death is decreased when ALS MuVs are added to the culture medium of cells deficient for FUS (FUS expression level was reduced by 79.5% ± 4.2% with siRNA strategy, 10 frames per well analysed, with n=3 wells per condition). **, P<0.01 ALS MuV treated cells respectively. ^TT^ P<0.01 ALS MuV treated scramble-RNA cells respectively. (F) FUS mRNA level is significantly higher in MN compared to myotubes. **, *P*<0.01 significantly different from human myotubes (n=3 per group). Values are means ± SEM. See Figure S5.

## Discussion

Extracellular vesicles are suspected to carry toxic elements from astrocytes towards motor neurons ^7^, and pathological proteins have recently been identified in circulating extracellular vesicles of sALS patients ^33^. Here, ALS muscle vesicles are shown to be toxic to motor neurons, which establishes the skeletal muscle as a potential source of vesicle-mediated toxicity in ALS.

The accumulation and over-secretion of muscle vesicles was observed as a consistent feature of sporadic ALS patients in this cohort, including patients carrying mutations in *C9orf72* or *ATXN2*, suggesting that this may be a common feature across many or all sporadic and familial forms of ALS. The consistent observation of a pronounced RNA processing and protein mislocalization phenotype in the ALS myotubes suggests that these cells recapitulate aspects of the disease mechanism that have been observed in motor neurons.

The observation that ALS MuVs act on RNA transport, that overexpression of wild-type FUS in MuV-recipient cells resulted in increased recipient cell death, and that RNA transport protein mislocalization is partially corrected and cell death reduced when FUS expression was knocked down in recipient cells, is consistent with a body of literature suggesting an RNA processing blockade mechanism in ALS MN ^34,35^. As shown, FUS expression is relatively high in iPSC MN compared to muscle cells (Figure 6F), and we also note that the cerebral cortex is among the tissues with the highest reported levels of FUS mRNA (http://www.proteinatlas.org ^36^). This could suggest a potential link between high expression levels of FUS in motor neurons and the susceptibility of recipient cells to the toxic contents of MuVs. It has previously been observed that motor neurons are selectively vulnerable to endocytosis of pathogenic proteins ^37,38^.

The observation that FUS and many of its binding partners are present in MuVs is of interest in the context of recent observations that normal function of FUS is required for normal neuromuscular junction (NMJ) development in mice, and that co-culture of iPSC-derived motor neurons with myotubes from FUS mutated patients resulted in impaired endplate maturation, which was proposed to be due to intrinsic FUS toxicity in both muscle and MN ^39^.

Several papers have described a muscle phenotype in ALS that occurs independently and prior to muscle denervation, such as metabolic imbalance ^40,41^, oxidative stress ^42^ and mitochondrial dysfunction ^43^. However, the role of muscle in ALS is unresolved. While an ALS-like phenotype was observed in mice when exogenous human mutant SOD1 expression was restricted to the skeletal muscle ^42,44^, other studies knocking down mutant SOD1 expression in skeletal murine muscle did not show any significant decreases in the progression of symptoms ^45,46^. The differences observed between these studies reveal the difficulties to assess the role of muscle in ALS, as the success of targeting whole skeletal muscle requires intravenous injection of a large amount of adeno-associated virus (AAV) particles ^47^. The secretion of muscle cell vesicles that are toxic towards motor neurons *in vitro* could suggest a potential role of muscle in ALS pathology. However, further *in vivo* experiments in appropriate animal models are required, to test the capacity of MuVs to diffuse *in vivo*, and their capacity to cross the blood brain barrier or to be absorbed directly at the neuromuscular junction.

## Data Availability

Associated large-scale datasets (transcriptomic and proteomic) are uploaded to public repositories. Expression data were uploaded to the GEO repository at accession number GSE122261. The mass spectrometry proteomics data have been deposited to the ProteomeXchange Consortium via the PRIDE partner repository with the dataset identifier PXD015736.

https://www.ncbi.nlm.nih.gov/geo/

https://www.ebi.ac.uk/pride/archive

## Acknowledgments

We thank the Human Cell Culture Platform of The Institute of Myology, and the “Plateforme Biopuces et Séquençage de l’IGBMC”. We thank Aswini Panigrahi for help with data upload. This work was financed by Target-ALS (ViTAL consortium, PI: S Duguez), ARsLA (TEAM consortium, PI: S Duguez ; PULSE, PI: D Devos), European Union Regional Development Fund (ERDF) EU Sustainable Competitiveness Programme for N. Ireland, Northern Ireland Public Health Agency (HSC R&D) & Ulster University (PI : T Bjourson). LLG is a recipient from ArSLA. It was also partially supported by European Community’s Health Seventh Framework Programme under grant agreement No. 259867 (Euro-MOTOR), INSERM, Sorbonne University, and the AFM. The study was sponsored by APHP.

The authors declare that they have no conflict of interest.

SD, LLG, SR, VMa, OC, VMi, EA, ZGO and UV performed the myoblast and motor neuron cell cultures and measurements, SK performed the nanosight, SK and KB performed the proteomic analysis on MuVs. JLGDA generated the transcriptome data on differentiated myoblasts. WJD performed the computational analysis, JL performed the electron microscopy analysis, SM performed genetic analysis. PB, AB, HB, GB, MDMA, DD, ACD, DFr, AH, AdHes, LL, PL, TL, PLeb, NLF, TM, VMe, AM, ZGO, FR, LR, FS, TS, GQ and PFP collected the muscle biopsies and clinical data the samples. The primary muscle myoblasts were subsequently extracted from the muscle biopsies by GO, SD, LLG. AlH generated the transcriptomic data. SD with the help of WJD took the lead in interpretation of results and writing the manuscript. CM, SK, JD, JLGDA, CR, GBB, OL discussed the results and provided critical feedback on the manuscript. SD and PFP conceived the original idea. SD supervised the project. The authors of this manuscript certify that they comply with the <u>ethical guidelines for authorship and publishing</u> in the Journal of Cachexia, Sarcopenia and Muscle^48^.

