## Supplemental Figures and Table S1 for "Muscle cells of sporadic ALS patients secrete neurotoxic vesicles"

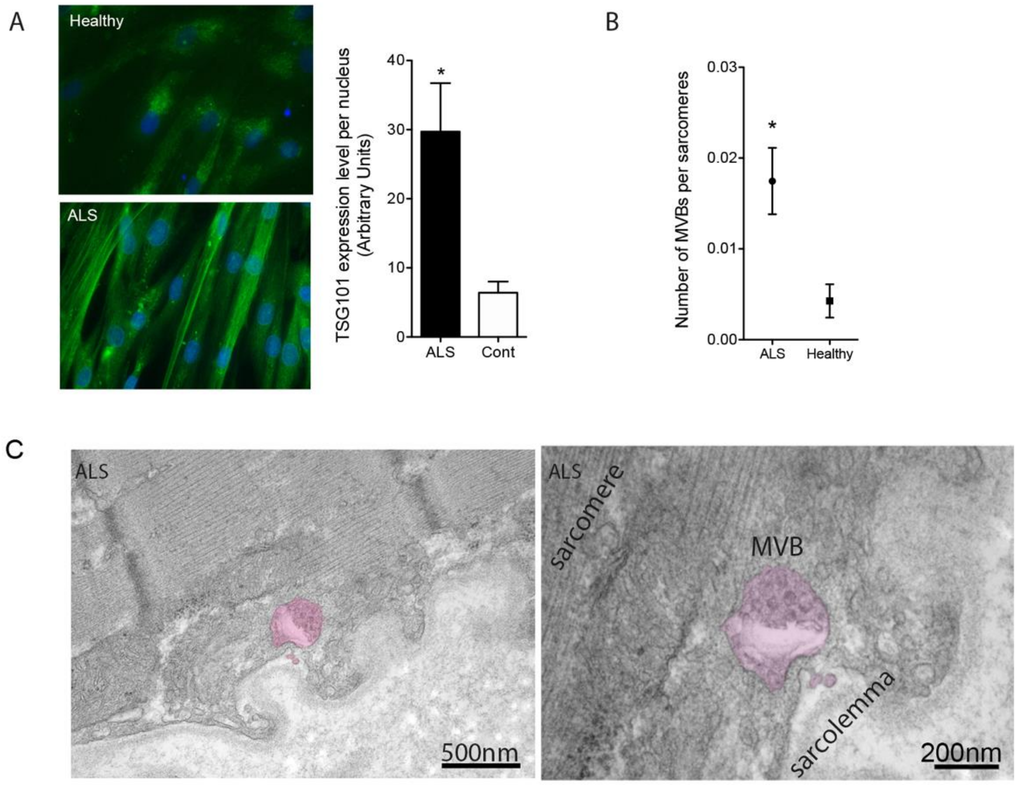


**Figure S1: Accumulation of exosomal markers in myotubes and muscle of ALS patients.** (A) Accumulation of TSG101 in ALS human myotubes. Left panel: representative images of TSG101 immunostaining performed on cultured myotubes from different patients. Right panel: Quantification of TSG101 fluorescence signal normalized per myonucleus (n=5 ALS and 6 healthy subjects). *, significantly different from healthy with P<0.05. (B) Quantification of multi-vesicular bodies per sarcomeres that are present in ALS and healthy muscle biopsies. n=3 ALS and 3 healthy subjects). *, significantly different from healthy with P<0.05. (C) Representative electron micrograph of sALS muscle longitudinal section. Multivesicular body (MVB) is highlighted in purple. The MVB contain exosome-like vesicles.

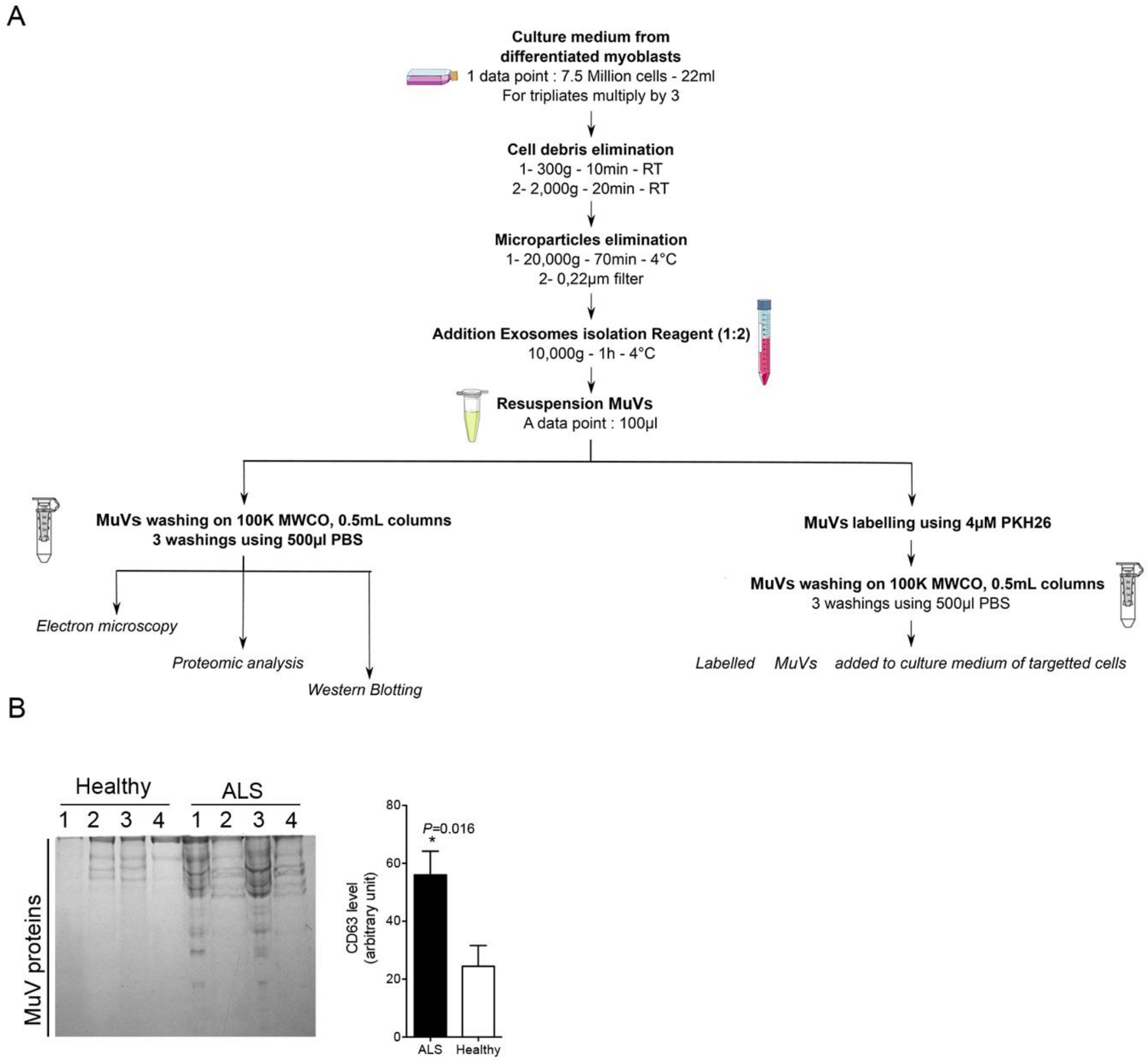


**Figure S2: More vesicles are secreted by ALS than by healthy myotubes**

(A) Muscle vesicle extraction protocol adapted for primary muscle stem cells. Conditioned media from differentiated myoblasts were cleared of cell debris, apoptotic bodies and microparticles through sequential centrifugations and filtration. Muscle vesicles were then precipitated using exosome isolation reagent and washed with PBS using 100k MWCO column. Rinsed MuVs were then either used for proteomic analysis and western blot, or labelled and added to the culture medium of targeted cells. (B) SDS-PAGE analysis showing a greater quantity of protein in muscle vesicles enriched fraction secreted by 800,000 ALS myotubes compared to healthy controls. Left panel: representative image of SDS-PAGE. Right panel: quantification of protein levels per well. *, significantly different from healthy, $P<0.05$.

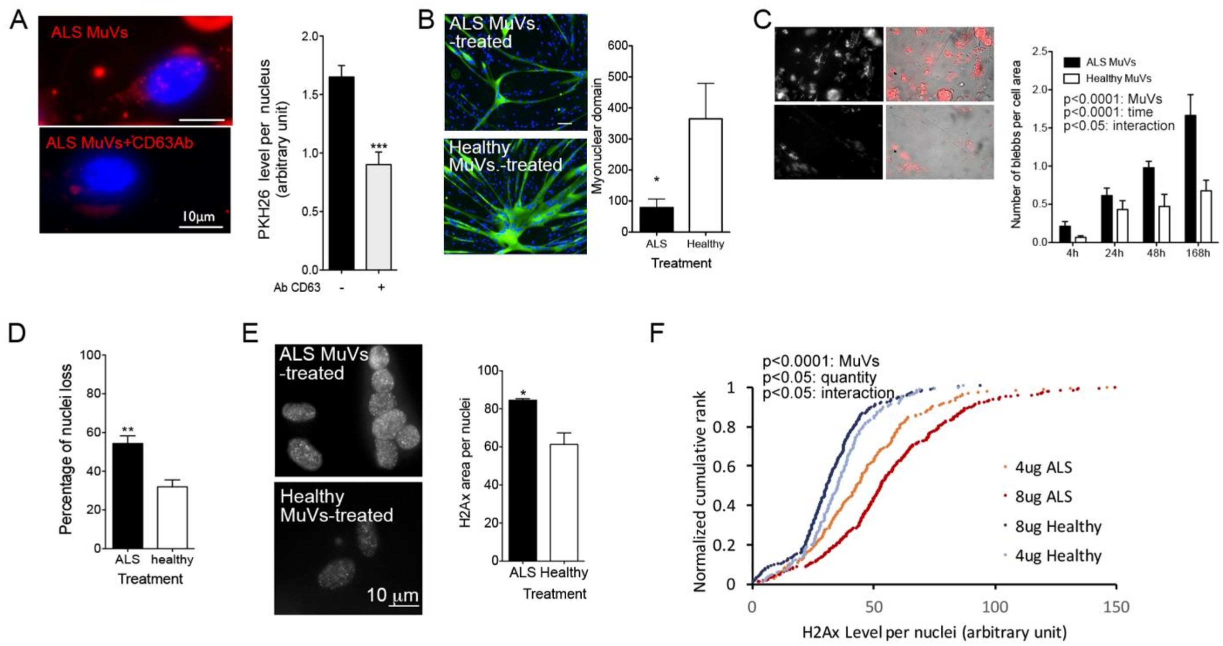


**Figure S3: ALS muscle vesicles are toxic toward healthy human myotubes.**

(A) Pre-treated the MuVs with CD63 antibody significantly decrease their uptake iPSC-MN. Left panel: representative images showing the ALS MuVs uptake by iPSC-MN. MuVs were labelled with PKH26. Right panel: quantification of PKH26 labelled MuVs on recipient cells. ***, P<0.001 versus non-CD63 pre-treated. (B) ALS MuVs induce muscle cell atrophy. Left panel: Representative images of healthy myotubes treated with ALS or Control MuVs. Myotubes are stained with myosin heavy chain (green). Right panel: quantification of myonuclear domain size (area of myosin heavy chain staining divided by the number of nuclei). The ALS-MuVs treated myotubes have a smaller myonuclear domain. * P<0.05, significantly different from healthy values. 2,000 to 4,000 myonuclei were analysed per subject, with n=3 subjects per group. (C) ALS MuVs induce cell stress. Counts of blebs per healthy myotube at different time-points after treatment with ALS or control MuVs. Five myotubes per time point per subject were analysed, with n=3 subjects per group. ANOVA 2 factor interaction, time and treatment P<0.001. (D) Levels of cell death marker H2Ax are increased in myonuclei of ALS-MuVs-treated myotubes compared to Healthy-MuVs-treated myotubes. Two hundred to 400 myonuclei per subject were analysed, with n=3 subjects per group. * P<0.05, significantly different from healthy values. (E) A greater loss of myonuclei is observed in cultures treated with ALS MuVs compared to healthy MuVs. n=3 subjects per group. ** P<0.01, significantly different from healthy controls. (F) Decreasing ALS MuVs quantity leads to decreased cell death. Left panel: representative immunostaining of H2-Ax immunostaining of healthy myotubes treated with ALS or control MuVs. Right panel: The graph represents a distribution of H2Ax level in myonuclei treated with either 4 or 8 mg of ALS MuVs (curves in orange and red), or with 4 or 8 mg of healthy MuVs (curves in light and dark blue). One hundred and eighty to 300 nuclei were analysed per subject and per condition, with n = 3 per subject. Two factor ANOVA was performed, showing the effect of ALS MuVs (P<0.0001), the effect varying due to quantity (P=0.0110), and an interaction between the two parameters (P=0.0301).

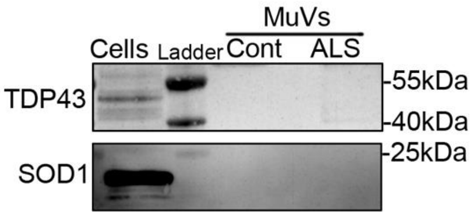


**Figure S4: Representative western blot showing the absence of detection of SOD1 and TDP43 in MuVs.**

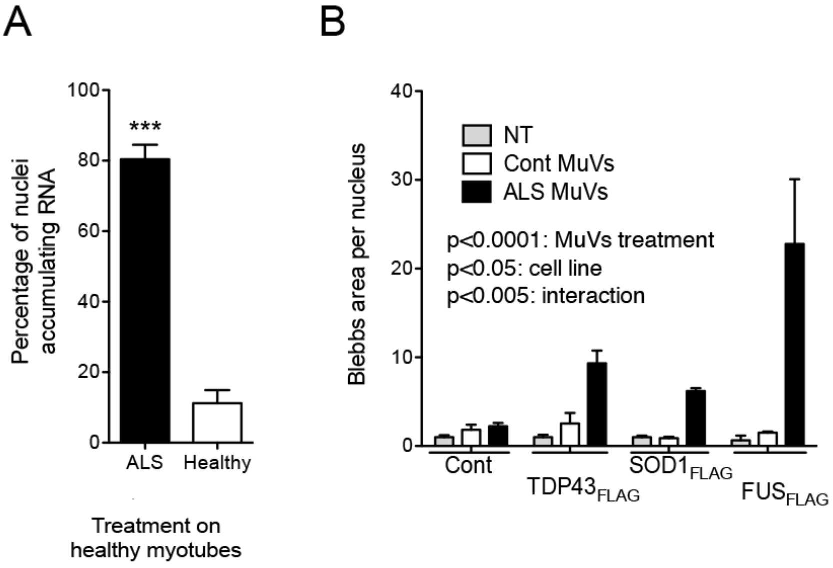

**Figure S5: ALS MuVs affect RNA transport in healthy myotubes and induce cell stress.**

(A) Percentage of myonuclei positive for accumulation of RNA in myotubes treated with ALS or healthy MuV. Accumulation of RNA is increased in the myonuclei of ALS-MuVs-treated myotubes. *** $P<0.001$, significantly different from healthy values. Seventy to 100 myonuclei per subject were analysed, with n=4 subjects per group. (B) Cell stress induced by ALS MuVs is exacerbated in presence of over-expression of FUS. ANOVA 2 factors was performed, showing the effect of ALS MuVs ($P<0.0001$), the effect varying due to cell line ($P<0.005$), and an interaction between the two parameters ($P<0.005$).

### Supplemental Tables

| ALS/healthy | 57 subjects | Age at time of biopsy | Gender | ALS/FRS | Genetics | Muscle testing | Site of onset 1=Upper limb 2=Lowe limb; 3=bulbar; 4=respiratory | Disease duration (months) | Histology | Electron microscopy | Cell culture | Transcriptome | Immunostaining cell culture | RT-qPCR | MuVs for proteomic analysis | MuVs for western blot | MuVs treatment MN | MuVs treatment myotubes |
|---|---|---|---|---|---|---|---|---|---|---|---|---|---|---|---|---|---|---|
| ALS | 27 ALS patients 8F, 19M 57.44±9.47 | 50-59 | F | 38 | - | 130 | 1 | 13 | x | | | | | | | | | |
| ALS | | 60-69 | M | 45 | - | 146 | 2 | 25.2 | | | x | x | | | | x | | x |
| ALS | | 50-59 | M | 41 | - | 140 | 2 | 26.7 | | | x | x | | | | | | x |
| ALS | | 50-59 | F | 39 | *C9orf72* | 146 | 3 | 16.5 | | | x | | x | | | x | x | |
| ALS | | 50-59 | F | 28 | *C9orf72* | 119 | 3 | 6 | x | | x | | x | x | | x | | x |
| ALS | | 60-69 | M | 38 | *ATXN2* | 130 | 2 | 11.3 | | | x | x | | x | | | x | |
| ALS | | 60-69 | M | 40 | - | 130 | 1 | 39.3 | | | x | x | | x | | x | | x |
| ALS | | 70-79 | M | 37 | - | 126 | 3 | 48.3 | | | x | x | | x | | x | | x |
| ALS | | 60-69 | M | 33 | - | 123 | 1 | 53.4 | x | | x | x | | x | | x | x | |
| ALS | | 70-79 | M | 32 | - | 104 | 1 | 28 | | | x | | x | | x | x | | |
| ALS | | 60-69 | F | 30 | - | 97 | 1 | 71.6 | | | x | | x | | | x | | |
| ALS | | 40-49 | M | 31 | - | 90 | 1 | 70 | | | x | | | | | x | | |
| ALS | | 50-59 | F | 23 | - | 103 | 3 | 33 | | | x | | | | | x | x | |
| ALS | | 60-69 | M | 40 | - | 97 | 1 | 11 | | | x | | x | | | x | | |
| ALS | | 60-69 | M | 38 | - | 88 | 1 | 52 | | | x | | x | | | x | | |
| ALS | | 50-59 | M | 31 | - | 94 | 2 | 40 | | | x | | | | | x | x | x |
| ALS | | 50-59 | M | 37 | - | No data | 0 | 101 | | x | | | | | | | | |
| ALS | | 30-39 | F | 32 | - | No data | 0 | 85 | | x | | | | | | | | |
| ALS | | 50-59 | F | 43 | - | 135 | 1 | 33.6 | x | | x | | x | | | x | | |
| ALS | | 40-49 | M | 41 | - | 111 | 2 | 44.1 | | | x | | x | x | | x | | x |
| ALS | | 50-59 | M | 40 | - | 130 | 2 | 17.4 | | | x | | x | x | | x | | |
| ALS | | 50-59 | M | 37 | *C9orf72* | 145 | 3 | 20.6 | | | x | | x | | x | x | | x |
| ALS | | 60-69 | M | 41 | - | 147 | 2 | 28.7 | | | x | | x | | | x | x | |
| ALS | | 50-59 | M | 36 | - | 139 | 2 | 12 | x | | | | | | | | | |
| ALS | | 60-69 | M | 43 | - | 149 | 1 | 12 | x | | x | | x | | | x | x | |
| ALS | | 30-39 | M | 41 | - | 144 | 1 | 12 | | | x | | x | | x | x | | |
| ALS | | 50-59 | F | 40 | - | 98 | 3 | 14 | | x | | | | | | | | |
| Healthy | 30 Healthy 10F ; 20M 51.08±18.40 | 60-69 | M | - | - | - | - | - | x | | | | | | | | | |
| Healthy | | 70-79 | M | - | - | - | - | - | x | | | | | | | | | |
| Healthy | | 50-59 | F | - | - | - | - | - | x | | | | | | | | | |
| Healthy | | 60-69 | F | - | - | - | - | - | x | | | | | | | | | |
| Healthy | | 50-59 | M | - | - | - | - | - | x | | | | | | | | | |
| Healthy | | 50-59 | F | - | - | - | - | - | | | x | | x | | | x | x | |
| Healthy | | 40-49 | F | - | - | - | - | - | | | x | | x | x | | x | | |
| Healthy | | 50-59 | M | - | - | - | - | - | | | x | | | | | x | x | |
| Healthy | | 40-49 | M | - | - | - | - | - | | | x | x | | | | x | | |
| Healthy | | 50-59 | M | - | - | - | - | - | | | x | x | | x | | x | x | |
| Healthy | | 50-59 | M | - | - | - | - | - | | | x | x | x | | | x | | x |
| Healthy | | 50-59 | F | - | - | - | - | - | | | x | | x | x | x | x | | x |
| Healthy | | 40-49 | F | - | - | - | - | - | | | x | | | x | | x | | x |
| Healthy | | 50-59 | M | - | - | - | - | - | | | x | x | x | x | | x | | x |
| Healthy | | 70-79 | M | - | - | - | - | - | | | x | x | | x | | x | x | |
| Healthy | | 20-29 | M | - | - | - | - | - | | | x | x | | | | | | |
| Healthy | | 20-29 | M | - | - | - | - | - | | | x | | | | | | | x |
| Healthy | | 50-59 | M | - | - | - | - | - | | x | | | | | | | | |
| Healthy | | 60-69 | F | - | - | - | - | - | | x | | | | | | | | |
| Healthy | | 60-69 | M | - | - | - | - | - | | x | | | | | | | | |
| Healthy | | 20-29 | M | - | - | - | - | - | | | x | | x | | | x | x | x |
| Healthy | | 30-39 | M | - | - | - | - | - | | | x | | x | | x | x | x | |
| Healthy | | 20-29 | M | - | - | - | - | - | | | x | | x | | x | x | x | x |
| Healthy | | 70-79 | M | - | - | - | - | - | | | x | | x | | | | | |
| Healthy | | 70-79 | F | - | - | - | - | - | | | x | | x | | | | | |
| Healthy | | 70-79 | M | - | - | - | - | - | | | x | | x | | | | | |
| Healthy | | 80-89 | M | - | - | - | - | - | | | x | | x | | | | | |
| Healthy | | 20-29 | M | - | - | - | - | - | | | x | | x | | | x | | |
| Healthy | | 60-69 | F | - | - | - | - | - | | | x | | x | | | x | | |
| Healthy | | 40-49 | F | - | - | - | - | - | | | x | | x | | | x | | |

**Table S1: Table summarizing the subject samples used for each type of experiment.**

Subject age range at time of biopsy is indicated in the column "Age". All ALS patients were confirmed to be ALS according to El Escorial. The ALSFRS-R and Muscle Testing results measured at the time of the biopsies are given. Manual muscle testing was scored from 0, representing total paralysis, to 5, representing normal strength, according to the Medical Research Council Score.

**Table S2: Table showing counts of peptides that were observed at consistently higher levels in exosomes of ALS myotubes compared to healthy controls.**

Peptides were filtered first to keep those for which the lowest value observed in ALS was greater than the highest value observed in controls, then were ranked by the difference in median values between ALS and controls.

**Table S3: Table showing counts of peptides that were observed in at least one ALS muscle exosome sample but not in any healthy controls.**
